# Commercialized kit to assess T-cell responses against SARS-CoV-2 S peptides. A pilot study in Health Care Workers

**DOI:** 10.1101/2021.03.31.21254472

**Authors:** Mónica Martínez-Gallo, Juliana Esperalba-Esquerra, Ricardo Pujol-Borrell, Víctor Sandá, Iria Arrese-Muñoz, Candela Fernández-Naval, Andrés Antón-Pagarolas, Victoria Cardona, Moisés Labrador-Horrillo, Tomás Pumarola-Suñé, Manuel Hernandéz-González

## Abstract

**Background:** It is crucial to assess the levels of protection generated by natural infection or SARS-CoV-2 vaccines, mainly in individuals professionally exposed and in vulnerable groups. Measuring T-cell responses may complement antibody tests currently in use as correlates of protection. Our aim was to assess the feasibility of a validated assay of T-cell responses.

**Methods:** Twenty health-care-workers (HCW) were included. Antibody test to SARS-CoV-2 N and S-proteins in parallel with a commercially available whole-blood-interferon-gamma-release-assay (IGRA) to S-peptides and two detection methods, CLIA and ELISA were determined.

**Results:** IGRA test detected T-cell responses in naturally exposed and vaccinated HCW already after first vaccination dose. The correlation by the two detection methods was very high (R>0.8) and sensitivity and specificity ranged between 100 and 86% and 100-73% respectively. Even though there was a very high concordance between specific antibody levels and the IGRA assay in the ability to detect immune response to SARS-CoV-2, there was a relatively low quantitative correlation. In the small group primed by natural infection, one vaccine dose was sufficient to reach immune response plateau. IGRA was positive in one, with Ig(S) antibody negative vaccinated immunosuppressed HCW illustrating another advantage of the IGRA-test.

**Conclusion:** Whole-blood-IGRA-tests amenable to automation and constitutes a promising additional tool for measuring the state of the immune response to SARS-CoV-2; they are applicable to large number of samples and may become a valuable correlate of protection to COVID-19, particularly for vulnerable groups at risk of being re-exposed to infection, as are health-care-workers.

## Introduction

One of the groups with the highest incidence of COVID-19 in the present pandemic has been Health Care Workers (HCW). In the last month of 2020, a worldwide vaccination campaign against SARS-CoV-2 began and, front line Health-Care-Workers (HCWs) have been among the first group receiving the new vaccines. As part of the adaptive response to infection, humans generate SARS-CoV-2-specific-antibodies and specific T-lymphocytes (1). Several studies on acute and convalescent COVID-19 patients have demonstrated that SARS-CoV-2-specific T cell responses control viral replication and reduce disease severity (2). Anti-spike (S) and anti-nucleocapsid (N) proteins IgG antibodies are associated with greatly reduced risk of SARS-CoV-2 reinfection in the 6 months after COVID-19 (3).

S-antigen-mRNA-based-vaccines against SARS-CoV2 (Pfizer-BioNTech and Moderna) have demonstrated immunogenicity (typically evaluated by serology) with 95% efficacy without major safety issues in all phases of human trials (4).). Although clinical trial data of vaccines in use are excellent in terms of effectiveness, real-world evidence remains scarce. One issue arisen in the health system -as in other essential sectors-is how to reassure HCWs, especially those pertaining to vulnerable groups e.g., diabetics, that, after overcoming COVID-19 or vaccination, they are reasonably protected and can reassume their duties.

Here we present a pilot study of SARS-CoV-2 specific antibody and T-cell responses to spike-protein (S) after mRNA-vaccination in a small group of HCWs at Hospital Universitari Vall d’Hebron (HUVH) using one commercially available test suitable for clinical laboratories, amenable to automation and therefore applicable to a large number of samples.

## Material and Methods

### Patients

Twenty HCWs of HUVH were recruited as part of a longitudinal study of seroprevalence and clinical impact of COVID-19 HUVH, a major academic hospital with over 6500 staff. All participants were tested before first dose (median 4 days, IQR 3), after first dose (median 22 days, IQR 1) and after the second dose (median 13 days, IQR 0) of the BNT162b2-mRNA-COVID-19-vaccine (Pfizer-BioNTech, Mainz, Germany). Demographic, epidemiological, and clinical data were collected using a standard questionnaire. Five had been diagnosed with COVID-19, (COVID group) based on clinical symptoms and positive PCR against SARS-CoV-2 and 15 remained unaffected (NO-COVID group).

This project was approved by the Hospital Universitari Vall d’Hebron Institutional Clinical Ethical Board. (HUVH PR(AG)113/2021).

## Methods

Antibody responses were measured in the clinical microbiology laboratory using two widely applied commercial CLIA assays; antibodies (IgG, IgM and IgA) to nucleocapsid (N) SARS-CoV-2 protein were detected by the qualitative *Elecsys*^*®*^ *Anti-SARS-CoV-2* test in a *Cobas*^*®*^ *8800 System* autoanalyzer (both from Roche Diagnostics, Mannheim, Germany); IgG antibodies to the Spike protein were measured by the quantitative LIAISON^®^ SARS-CoV-2 Trimeric S IgG test in a XL Analyzer (DiaSorin, Stillwater, MN, USA). Samples with antibody levels >800 UA/mL were diluted according to the manufacturer’s instructions.

SARS-CoV-2-specific T-cell responses were assessed in the clinical immunology laboratory by a whole blood Interferon-Gamma-Release-immuno-Assay (IGRA) that uses two Qiagen^®^ (Hilden, Germany) proprietary mixes of SARS-CoV-2 S-protein (Ag1 and Ag2) selected to activate both CD4 and CD8 T-cells, following manufacturer’s instructions. Briefly, venous blood samples were collected directly into the Quantiferon^®^ tubes containing spike peptides as well as positive and negative controls. Whole blood was incubated at 37°C for 16-24 hours and centrifuged to separate plasma. IFN-γ (IU/ml) was measured in these plasma samples in parallel using CLIA (Liason, Quantiferon^®^ Gold Plus) and ELISA (QuantiFERON^®^ Human IFN-γ SARS-CoV-2, Qiagen^®^) tests, both for research only use. Complete blood Count (CBC), flow cytometry lymphocyte phenotype and immunoglobulin levels were obtained in parallel samples (5,6).

### Statistical analysis

Data were analyzed by non-parametric tests; Mann-Whitney test for comparisons between the COVID and NO-COVID groups, and Friedman test for comparisons between paired values, and correlation studies to compare variables was calculated by Pearson correlation coefficient. GraphPad Prism v8.01 software was used for both statistical analysis and graphical representation.

## Results

### In the COVID group plateau IgG (S) antibody levels were attained with first vaccination dose

At enrolment COVID group participants (n=5) had detectable IgG (N) antibodies while those in the NO-COVID group, (n=15) were all negative, thus excluding the presence of asymptomatic cases in the latter group (Figure 1A). IgG(S) antibodies were measured at three time points: prior and after the vaccine first dose and after the boost second dose. NO-COVID group participants were all negative for IgG(S) antibodies. After first vaccination, all NO-COVID subjects but one, developed IgG(S) antibodies although not reaching average levels to those found in the COVID group prior to vaccination (Mean (STDV) 150.3 (99.8) vs 2712 (511) respectively). The boost vaccine dose induced in them a further increase in IgG(S) antibody levels but not in the COVID group (Figure 1B) as if they had reached their plateau (see also Figure 2).

**Figure 1.**
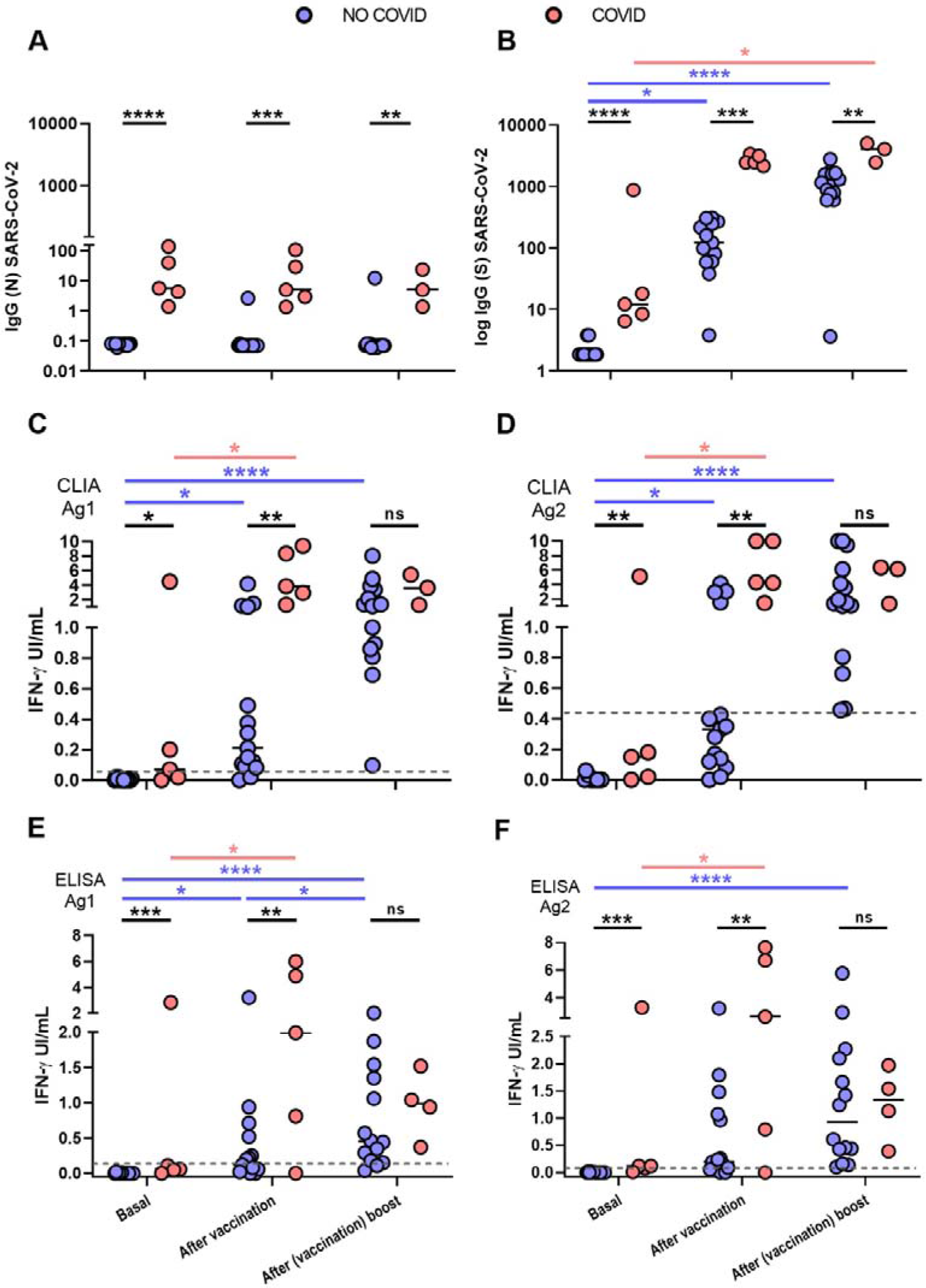
Antibody and cellular immune responses to SARS-CoV-2 in the two groups of HCW, NO-COVID (blue) and COVID (pink) after BNT162b2-mRNA-COVID-19 vaccination at three time points: before 1st dose, before 2nd dose and after 2nd dose. A and B, results of IgG antibody levels to N (A) and S SARS-CoV-2 (B) proteins. C to F. T cells responses to SARS-CoV-2 S peptides in two IGRA tests that measure IFN-gamma production by CLIA (A & B) and ELISA (E & F). Significance of the differences, *p<0.05, **p<0.01, ***p<0.001.****p<0.0001.

**Figure 2.**
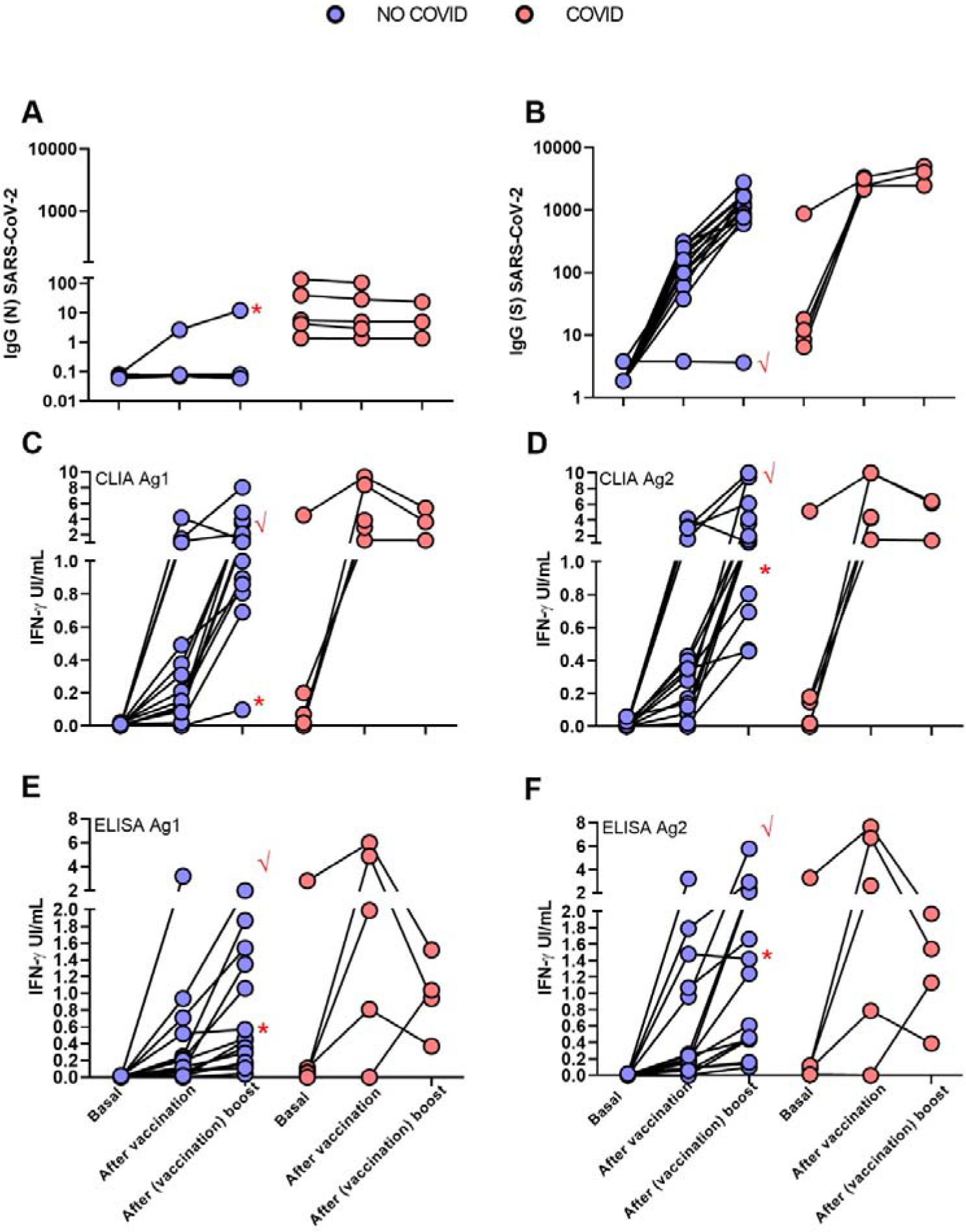
Follow up of antibody and cellular immune responses to SARS-CoV-2 in the two groups of HCW, NO-COVID (blue) and COVID (pink) after BNT162b2-mRNA-COVID-19 vaccination at three time points: before 1st dose, before 2nd dose and after 2nd dose. A and B, results of IgG antibody levels to N (A) and S SARS-CoV-2 (B) proteins. C to F. T cells responses to SARS-CoV-2 S peptides measuring IFN-gamma production by two detection methods: CLIA (A & B) and ELISA (E & F). Patient 12 (√) and patient 19 (*) are labelled by red symbols.

### IGRA detects specific T-cell response to SARS-CoV-2 with high sensitivity and specificity using either CLIA or ELISA tests to measure IFN-γ production

Results from IFN-γ production to Ag1 and Ag2 S-peptides measured by CLIA are in Figure 1C and D and by ELISA in E and F. The correlation of CLIA and ELISA readings was almost total (R>0.9) in all cases.

Cut-off points for each variant of the IGRA assay were calculated by ROC analysis with the following results: CLIA cut-off point was > 0.051 IU IFN-gamma/ml with a sensitivity of 86.6% and specificity of 100% for Ag1 and > 0.44 IU IFN-gamma/ml for Ag2 with a sensitivity 100% and a specificity 73.3% (Figure 3). The cut-off point for the ELISA was >0.13 IU IFN-gamma/ml for Ag1 with a sensitivity of 85.7% and specificity 100% and for Ag2 >0.12 IU IFN-gamma/ml with a sensitivity of 92.8% and a specificity of 100%. Correlation between Ag1 and Ag2 results was very high (R=0.88 for ELISA and 0.84 for CLIA (data not shown). In general Ag2 induced higher responses than Ag1, and. A positive response to either of the two peptides pools should be considered, in principle a positive result. From these results, it is not clear the need of using the Ag1 pool of peptides but this may be different in a larger group with a more diverse genetic background.

**Figure 3.**
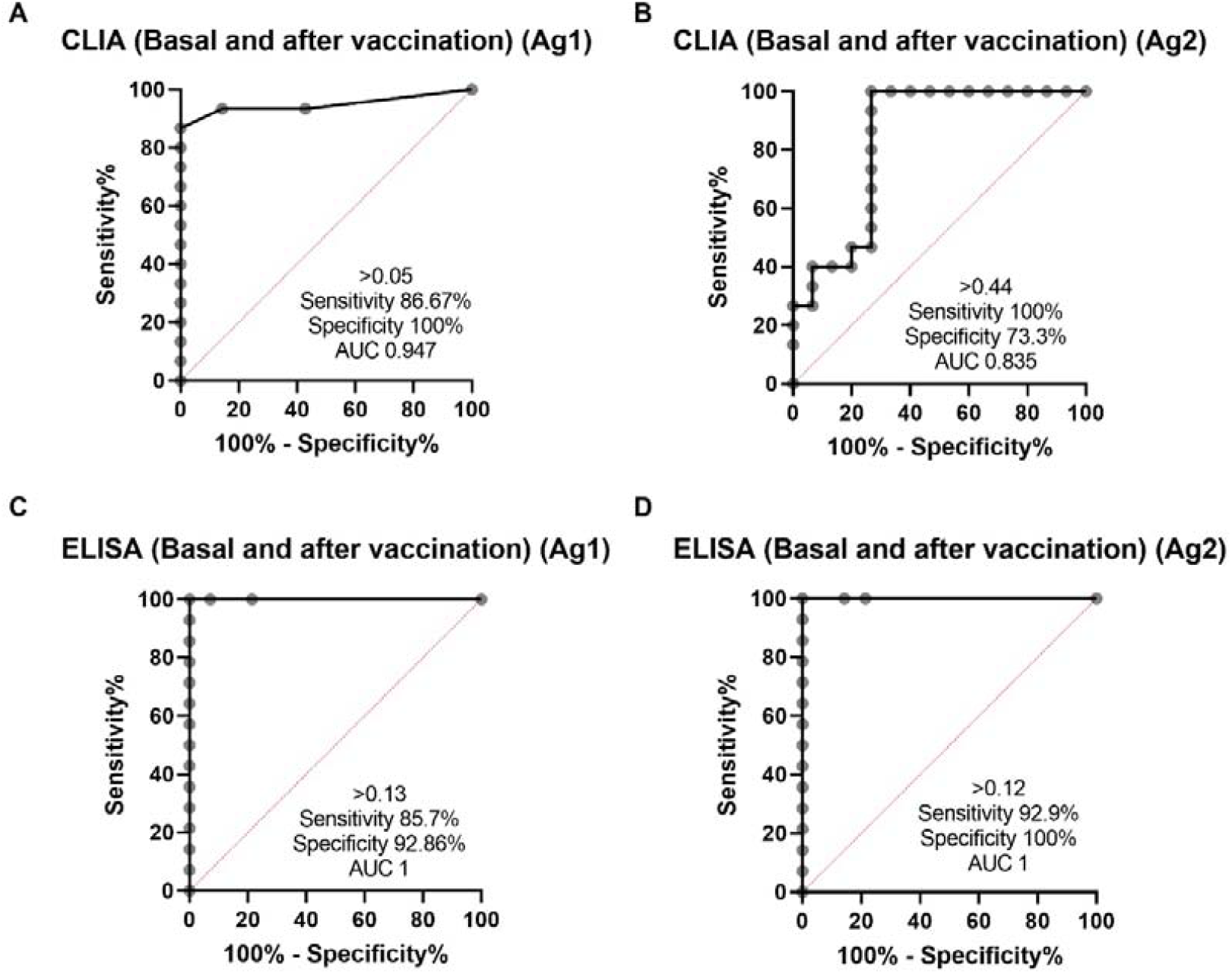
Analysis of the methods for assessing cell-mediated immunity by SARS-CoV-2 IGRA-type CLIA (A and B) and ELISA (C and D) using ROC curves comparing basal and after vaccine study points in NO-COVID group.

Overall, quantitative correlation between the antibody and T-cell responses was low (R in the order of 0.2) (Figure 4) but there was a good concordance in detecting the immune response to Spike. Sensitivity of the IGRA test may be superior in this group to IgG(S) antibody test due to the presence of one anti-CD20 treated participant (see below, illustrative case 12), but due to the small size, conclusions cannot be drawn.

**Figure 4.**
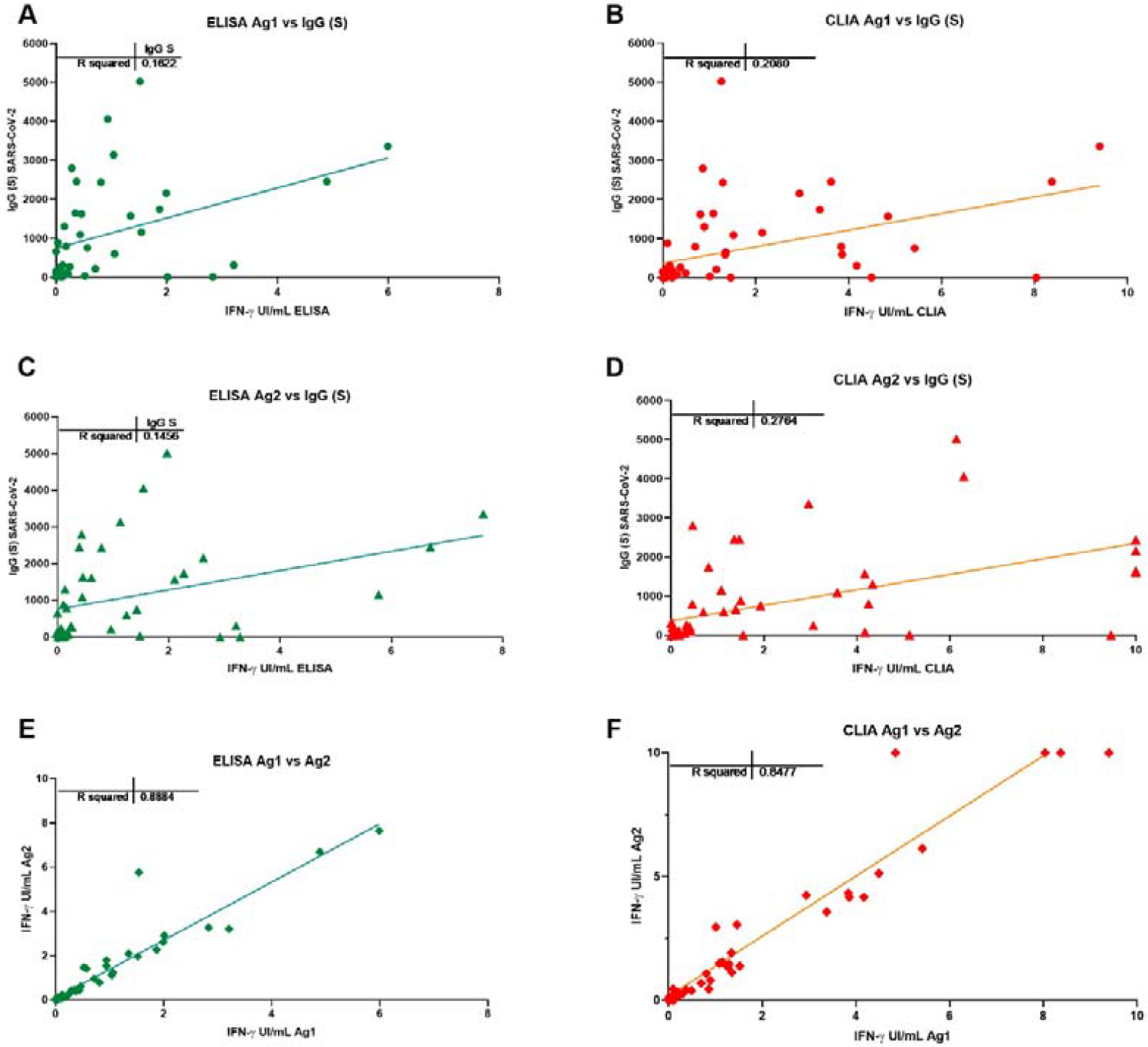
Correlation between the antibody and T cell responses with different IFN-g detection methods ELISA (A and C) and CLIA (B and D). Correlation between the T cell response to Ag1 and Ag2 with ELISA (E) and CLIA (F).

### One vaccine dose restores T cell response in post COVID participants, but vaccination boost was required for naïve participants to attain a good response

Interestingly four of the five post COVID group did not respond in the IGRA test prior to vaccination, but one dose was sufficient to reach good levels of IFN-γ production, indicating priming by the natural infection. Second dose rather reduced the response, perhaps because due to the time elapsed since priming, response contracted more quickly, this reduction has now been reported in two not yet peer reviewed reports (7, 8). In the NO-COVID group even if the first dose already induced a significant response, the boost vaccination was required to reach a response like that of the COVID group after first dose.

In this small study, we did not find any relation of antibody nor T-cell responses to spike proteins with the total lymphocytes, their main subsets, or the immunoglobulin levels. There was a trend for antibody and T-cell responses to be lower in older patients (Figure 5), no gender effect was detected.

**Figure 5.**
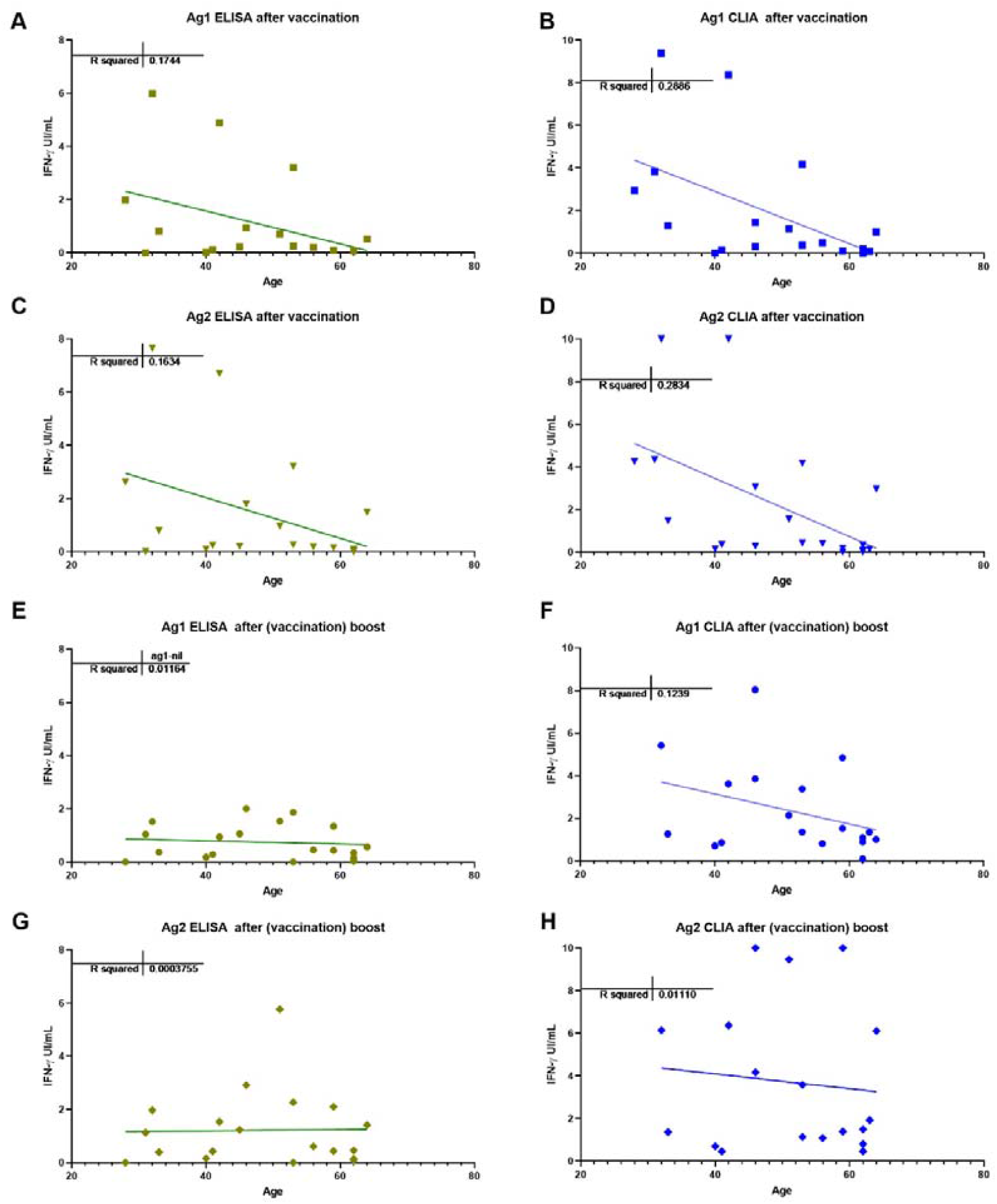
Correlation between age and T cell responses with different IFN-g detection methods ELISA (A, C, E and G) and CLIA (B, D, F and H) with Ag1 and Ag2; after vaccine (A, B, C and D) and after (vaccine) boost (E, F, G and H).

### Illustrative HCW cases

As mentioned above one of the NO-COVID patients, case 12, was treated with anti-CD20 for an autoimmune condition and did not develop IgG or IgM antibodies after vaccination. Interestingly a clearly measurable T-cell responses was detected (see Figure 2 tick case) after vaccination and a small expansion of circulating of plasmablasts in sequential samples. We detected plasmablast after the vaccine boost with few pre class switch B lymphocytes prior to vaccination despite anti-CD20 treatment, indicating residual capacity of the B-cell compartment (Figure 6).

**Figure 6.**
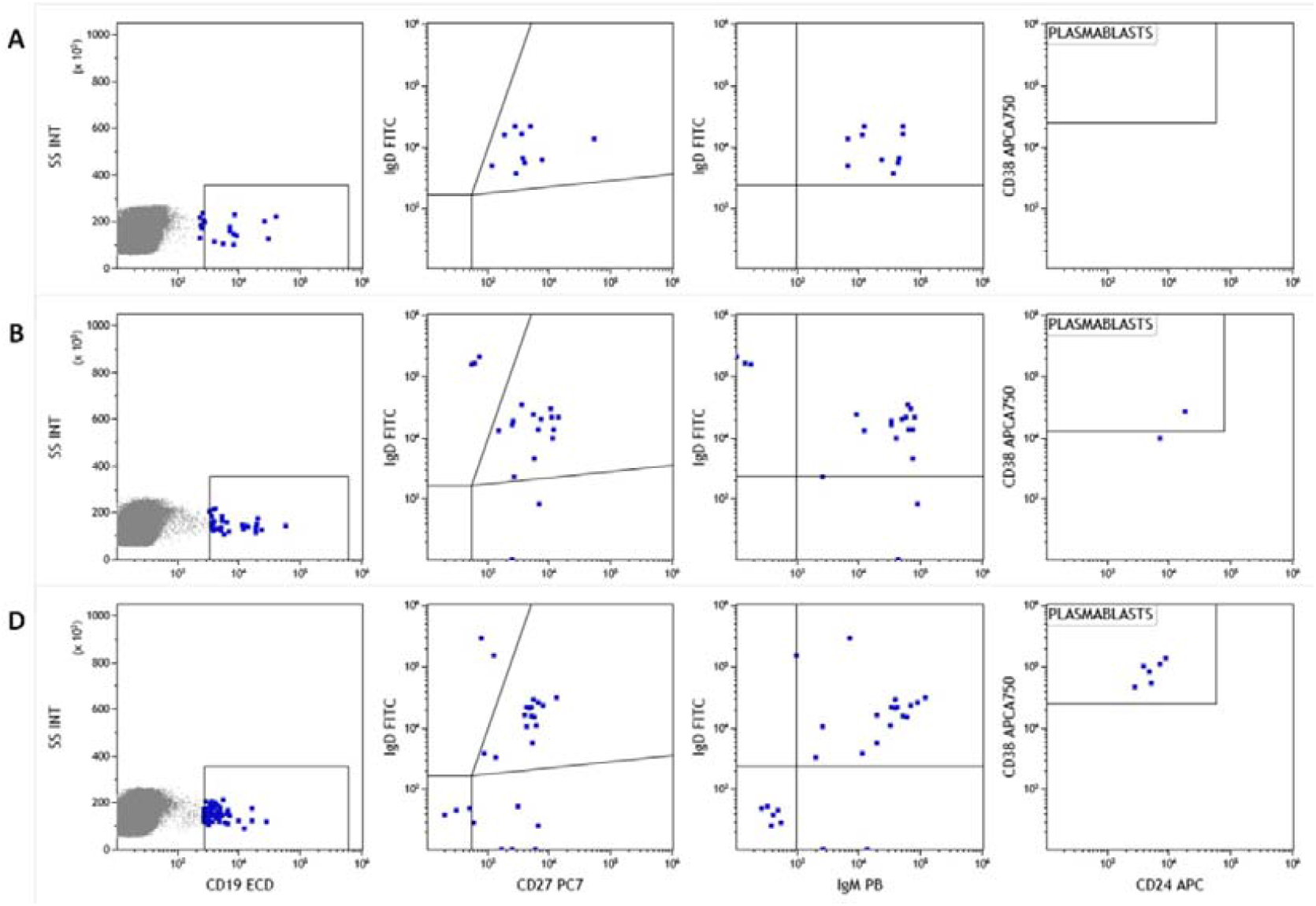
Analysis of naive B lymphocyte subpopulations in patient 12. Class switch, IgM memory B cell populations and plasmablasts before vaccination (A), after first dose (B) and second vaccination dose (C) Memory/switch B-lymphocytes were already present before vaccination and plasmablasts appeared only after vaccination.

Another NO-COVID group patient, case 19, was infected the same day that she received first dose of vaccination. The patient developed T-cell response in the IGRA-assay, and in this case, in addition to antibodies to the S antigen, she developed antibodies to N-antigen (Figure 1A, dot with asterisk), but the response was like the NO-COVID group indicating that being primed by two simultaneous stimuli did not result in this case in a supranormal response.

## Discussion

We have studied in parallel antibody and cellular response to the BNT162b2-mRNA-COVID-19-vaccine in a small group of HCW and found that the IGRA-assays yields rapid results that are concordant with antibody tests and could constitute a valuable contribution to the evaluation of the immune response in people that may need to be reassured of being protected against SARS-CoV2 infection. This is however, a small pilot study that should be expanded to really demonstrate that SARS-CoV-2 spike IGRA based tests can constitute valuable correlates of protection complementary to serology.

It has already been shown that protein N is the most immunogenic structural protein and that IgG responses to protein-S in SARS-CoV-2 are lower, possibly due to the glycosylation state of this protein (9). This may explain that IGRA and IgG(S) antibody tests were negative in two of the participants in the COVID group that were IgG(N) positive. However, after immunization with a single dose, they responded to a level higher than fully vaccinated NO-COVID participants.

There are a good number of studies demonstrating T-cell responses to SARS-CoV-2 using techniques (2) widely applied to measure responses to other viral infection (10,11). These techniques include intracellular-cytokine-staining (ICS) by flow cytometry and variations of the ELISPOT-assay which measure mainly IFN-γ production. Activation-induced markers (AIM), also by flow cytometry, can be a sensitive technique (12). However, they have limitations in the applicability compared with whole blood IGRA-assays used here when it comes to using them in a clinical laboratory (13).

Availability of a complementary correlate of protection in addition to serology may be invaluable for two groups, 1) HCW and other professionals with vulnerability factors that need to be reassured of being immunized against SARS-CoV-2 before reassuming tasks that have a risk of accidental re-exposure, and 2) for immunosuppressed patients that fail to make a measurable antibody response. IGRA spike peptides test may constitute a very valuable tool in this context as it can be applied to a large number of samples producing results in 24h and with promising sensitivity and specificity.

## Data Availability

27-March-2021

## Abbreviations

(HCWs): Health care workers
(IGRA): Interferon-gamma-release-assays
(CLIA): Chemiluminescence immunoassays
(ELISA): Enzyme linked immunosorbent assay
(CBC): Complete blood count
(S): Spike protein
(N): Nucleocapsid protein

## ACKNOWLEDGEMENTS

The authors want to express their gratitude to participants of the study and laboratory technicians from Microbiology and Immunology for technical assistance. We thank Qiagen and Diasorin^®^ for commercial kits donation, these companies do not participate in the experimental plan, data analysis nor in writing this manuscript. English writing support was provided by Fidelma Greaves.

This study was funded in part by Instituto de Salud Carlos III, grant COV20/00416 co-financed by the European Regional Development Fund (ERDF).

## Author disclosure statement

The authors declare that they have no conflict of interest.

All authors have accepted responsibility for the entire content of this manuscript and approved its submission.

V.S.M. I.A.M. and C.F.N. participated in performing the research and data analysis.

R.P.B., V.C., A.A.P., M.H.G. and T.M.S participate in critical review of the manuscript.

M.M.G. J.E.E. and M.L.H. participated in conducting the study, writing and reviewing the manuscript.

## Bibliography

1. Stephens DS, McElrath MJ. COVID-19 and the Path to Immunity. JAMA - Journal of the American Medical Association. 2020.

2. Rydyznski Moderbacher C, Ramirez SI, Dan JM, Grifoni A, Hastie KM, Weiskopf D, et al. Antigen-Specific Adaptive Immunity to SARS-CoV-2 in Acute COVID-19 and Associations with Age and Disease Severity. Cell. 2020;

3. Lumley SF, O’Donnell D, Stoesser NE, Matthews PC, Howarth A, Hatch SB, et al. Antibody Status and Incidence of SARS-CoV-2 Infection in Health Care Workers. N Engl J Med. 2021;

4. S A Meo, I A Bukhari, J Akram, A S Meo, D C Klonoff. COVID-19 vaccines: comparison of biological, pharmacological characteristics and adverse effects of Pfizer/BioNTech and Moderna Vaccines. Eur Rev Med Pharmacol Sci 2021.

5. Garcia-Prat M, Álvarez-Sierra D, Aguiló-Cucurull A, Salgado-Perandrés S, Briongos-Sebastian S, Franco-Jarava C, et al. Extended immunophenotyping reference values in a healthy pediatric population. Cytom Part B - Clin Cytom. 2019;

6. Garcia-Prat M, Vila-Pijoan G, Martos Gutierrez S, Gala Yerga G, García Guantes E, Martínez-Gallo M, et al. Age-specific pediatric reference ranges for immunoglobulins and complement proteins on the Optilite™ automated turbidimetric analyzer. J Clin Lab Anal. 2018;

7. Carmen Camara, Daniel Lozano-Ojalvo, Eduardo Lopez-Granados, Estela Paz-Artal, Marjorie Pion, Jordi Ochando et al. Differential effects of the second SARS-CoV-2 mRNA vaccine dose on T cell immunity in naïve and COVID-19 recovered individuals.

8. Adrienn Angyal, Stephanie Longet2, Shona Moore3, Rebecca P. Payne, Thushan I. de Silva et al. T-cell and antibody responses to first BNT162b2 vaccine dose in previously SARS-CoV-2-infected and infection-naive UK healthcare workers: a multicentre, prospective, observational cohort study.

9. Grant OC, Montgomery D, Ito K, Woods RJ. Analysis of the SARS-CoV-2 spike protein glycan shield reveals implications for immune recognition. Sci Rep. 2020.

10. Mario Fernández-Ruiz 1, Beatriz Olea, Estela Giménez, Rocío Laguna-Goya, Hernando Trujillo, Fernando Caravaca-Fontán, José María Aguado et al. SARS-CoV-2-Specific Cell-Mediated Immunity in Kidney Transplant Recipients Recovered from COVID-19. Transplantation. 2021

11. Julie Demaret, Guillaume Lefèvre, Fanny Vuotto, Jacques Trauet, Alain Duhamel, Myriam Labalette et al. Severe SARS-CoV-2 patients develop a higher specific T-cell response. Clin Transl Immunology. 2020

12. Grifoni A, Weiskopf D, Ramirez SI, Mateus J, Dan JM, Moderbacher CR, et al. Targets of T Cell Responses to SARS-CoV-2 Coronavirus in Humans with COVID-19 Disease and Unexposed Individuals. Cell. 2020;

13. Linda Petrone, Elisa Petruccioli, Valentina Vanini, Gilda Cuzzi, Saeid Najafi Fard, Delia Goletti et al. A whole blood test to measure SARS-CoV-2-specific response in COVID-19 patients. Clin Microbiol Infect. 2021

